# Low albumin levels are associated with poorer outcomes in a case series of COVID-19 patients in Spain: a retrospective cohort study

**DOI:** 10.1101/2020.05.07.20094987

**Authors:** Roberto de la Rica, Marcio Borges, María Aranda, Alberto del Castillo, Antonia Socias, Antoni Payeras, Gemma Rialp, Lorenzo Socias, Lluis Masmiquel, Marta Gonzalez-Freire

## Abstract

**OBJECTIVE:** To describe the clinical characteristics and epidemiological features of severe (non-ICU) and critically patients (ICU) with COVID-19 at triage, prior hospitalization, in one of the main hospitals in The Balearic Islands health care system.

**DESIGN:** Retrospective observational study

**SETTING:** Son Llatzer University Hospital in Palma de Mallorca (Spain)

**PARTICIPANTS:** Among a cohort of 52 hospitalized patients as of 31 March 2020, 48 with complete demographic information and severe acute respiratory syndrome coronavirus 2 (SARS-CoV-2) positive test, were analyzed. Data were collected between March 15th, 2020, and March 31th 2020, inclusive of these dates.

**MAIN OUTCOMES:** Clinical, vital signs and routine laboratory outcomes at the time of hospitalization, including symptoms reported prior to hospitalization. Demographics and baseline comorbidities were also collected. Mortality was reported at the end of the study.

**RESULTS:** 48 patients (27 non-ICU and 21 ICU) resident in Mallorca, Spain (mean age, 66 years, [range, 33-88 years]; 67% males) with positive SARS-CoV-2 infection were analyzed. There were no differences in age or sex among groups (p >.05). Initial symptoms included fever (100%), coughing (85%), dyspnea (76%), diarrhea (42%) and asthenia (21%). The majority of patients in this case series were hospitalized because of low SpO2 (SpO2 below 90%) and presentation of bilateral pneumonia (94%) at triage. ICU patients had a higher prevalence of dyspnea compared to non-ICU patients (95% vs 61%, p = .022). Acute respiratory syndrome (ARDS) was presented in 100% of the ICU-patients. All the patients included in the study required oxygen therapy. ICU-patients had lymphopenia as well as hypoalbuminemia. Inflammatory markers such as lactate dehydrogenase (LDH), C-reactive protein (CRP), and procalcitonin were significantly higher in ICU patients compared to non-ICU (p < .001).Lower albumin levels were associated with poor prognosis measured as longer hospital length (r= −0.472, p <.001) and mortality (r= −0.424, p=.003). Interestingly we also found, that MCV was lower among of those patients who died (p=.0002). As of April 28, 2020, 10 patients (8 ICU and 2 non-ICU) had died (21% mortality) and while 100% of the non-ICU patients had been discharged, 33% of ICU patients still remained hospitalized (5 in ICU and 2 had been transferred to ward).

**CONCLUSION:** Critically ill patients with COVID-19 present lymphopenia, hypoalbuminemia as well high levels of inflammation. Lower levels of albumin were associated with poorer outcomes in COVID-19 patients. Albumin might be of importance because of its association with disease severity in patients infected with SARS-CoV-2.

**WHAT IS ALREADY KNOWN IN THIS TOPIC:** Spain has been hit particularly hard by the pandemic. By the time that this manuscript was written more than 25.000 deaths related to COVID-19 have been confirmed. There is limited information available describing the clinical and epidemiological features of Spanish patients requiring hospitalization for COVID-19. Also, it is important to know the characteristics of the hospitalized patients who become critically ill

**WHAT THIS STUDY ADDS:** This small case series provides the first steps towards a comprehensive clinical characterization of severe and critical COVID-19 adult patients in Spain. The overall mortality in our patients was 21%. To our knowledge this is the first report with reporting these features in Spain. At triage the majority of patients had lower SpO2 (<90%) and bilateral pneumonia. The most common comorbidities were hypertension (70%), dyslipidemia (62%) and cardiovascular disease (30%). Critically ill patients present hypoalbuminemia and lymphopenia, as well as higher levels of inflammation. Albumin might be of importance because of its association with disease severity and mortality in patients infected with SARS-CoV-2.

## Introduction

The SARS-CoV-2 outbreak that originated in Wuhan in December 2019 has rapidly spread worldwide.^1^ Spain has been hit particularly hard by the pandemic. By the time that this manuscript was written more than 23.000 deaths related to COVID-19 have been confirmed. There is an urgent need to understand the causes behind these poor outcomes in order to improve patient management .^1-5^ Thus, it is imperative to clinically characterize critically ill COVID-19 patients in order to identify those with a bad prognosis at an early stage, before their situation becomes irreversible.

COVID-19 has a rather heterogenous presentation. While many patients remain asymptomatic carriers, others can show a wide array of symptoms, from mild flu-like manifestations such as dry cough, phlegm, myalgia or diarrhea to severe pneumonia or even acute respiratory distress syndrome (ARDS).^1,4^ The exact pathobiology responsible for severe and critically ill cases is still not clear. It has been proposed that a hyperinflammatory syndrome may play a central role in the progression from mild to severe or critical COVID-19.^6-10^ Inflammatory factors are likely involved in this process and could become biomarkers of disease progression in the near future.^11,12^ Judging from similar hyperinflammatory syndromes like bacterial sepsis, fluctuations of these biomarkers will probably be strongly interrelated and time dependent.^13^ Until this process is fully characterized, biochemical parameters and physical examinations are the only tools available for tracking disease progression. Efforts are being made in order to fully characterize the clinical characteristics of COVID-19 nationwide. The objective of this retrospective case series study was to describe the epidemiological and clinical characteristics of 48 hospitalized patients with COVID-19 and to compare patients who were admitted to the intensive care unit (ICU) care with those who did not receive ICU care, staying at ward.

## Methods

### Study population

The study was conducted at Son Llatzer University Hospital, a public tertiary care center covering 280.000 population from urban and rural areas in Mallorca, Balearic Islands, in Spain. The institutional review board approved this case series as minimal-risk research using data collected for routine clinical practice and waived the requirement for informed consent. According to the WHO guidance,^14^ laboratory confirmation for SARS-Cov-2 was defined as a positive result of real time reverse transcriptase–polymerase chain reaction (RT-PCR) assay from nasal and pharyngeal swabs. Patients with confirmed SARS-CoV-2 infection by positive result on the RT-PCR or serological test as of March 31, 2020 were admitted in the study. 52 patients met the inclusion criteria, but of those, only 48 had epidemiological data. Tests were repeated on inpatients presenting clinical signs of COVID-19 disease during hospitalization if the initial test was negative, likely to be a false-negative, or due to poor sample collection. Patients were admitted to Son Llatzer University Hospital between March 15, 2020, and March 31, 2020, inclusive of those dates. Clinical outcomes were monitored until the final date of follow-up. The follow-up data on clinical and laboratory measures are not included in this study.

### Data collection

Clinical and laboratory data were collected at triage by hospital staff (nurses). The data were recorded on electronic worksheets and uploaded to the health database. Three researchers independently reviewed the data collection forms for accuracy. Data collected included patient demographic information (age, sex, race, home medications, smoking habits), comorbidities, initial symptoms of the disease, triage vitals such as fever, oxygen saturation (SpO_2_), systolic and diastolic pressure, heart rate, as well as diagnosis of pneumonia by chest X-ray. In some instances, patients had missing data for the above parameters, in which case percentages of total patients with completed tests are shown. Initial laboratory testing was defined as the first test results available, typically within 24 hours of admission. Complete blood cell count, tests of kidney and liver function, and inflammatory and coagulation markers such as C-reactive protein, lactate dehydrogenase, D-dimer, fibrinogen, troponin I and procalcitonin levels were performed. Respiratory samples were tested for influenza and other respiratory viruses with a multiplex PCR assay. Patients underwent chest x-rays or computed tomography for pneumonia diagnosis. Supplemental oxygen was administered when saturations as measured by pulse oximeter dropped below 92%. Patients received antibiotics, anti-malaria drug (Hidroxychoroquine or Chloroquine), a co-formulated (“Kaletra”) antivirals lopinavir-ritonavir, immunosuppressive drugs such as Tocilizumab and Interferon beta, and anti-inflammatory drugs. All patients received oxygen therapy.

### Statistical Analysis

No statistical sample size calculation was performed a priori, and sample size was equal to the number of patients treated during the study period. Baseline characteristics of non-ICU versus ICU patients were summarized as means and SDs for continuous variables and as frequencies and percentages for categorical variables. T tests were used to compare continuous characteristics and Fisher exact tests were used to compare categorical characteristics of non-ICU versus ICU patients when appropriate. Non-normal distributed continuous data were compared using the Mann-Whitney-Wilcoxon test. Log scale for some variables are presented when “physiological” outliers were detected. In some cases, differences in clinical and laboratory measures between non-ICU and ICU patients were assessed in multivariable linear regression models adjusted for age, sex, race, smoking status, SpO_2_ and comorbidities. Relationships were assessed using spearman or Pearson correlations. All statistical tests were 2-tailed, and statistical significance was defined as p < .05. All analyses were performed using version 3.5.2 of the R programming language (R Project for Statistical Computing; R Foundation).

### Study approval

The study was performed in accordance with Good Clinical Practice and the Declaration of Helsinki principles for ethical research. Ethical approval for this project (IB 4165/20 PI) was obtained from the ethics committee of the Balearic Islands. Written informed consent was waived due to the rapid emergence of this infectious disease.

## Results

### Clinical features

In this retrospective study, the clinical and epidemiological characteristics of 48 patients (mean age, 66 years, [range, 33-88 years]; 67% males) with COVID-19 were analyzed. The patients were classified as non-ICU and ICU according to the severity and the guidelines of the Son LLatzer University Hospital for COVID-19 management. Table 1 shows the demographic, vital, and clinical characteristics of the patients. There were no differences in age or sex among groups (*p* >.05)Comorbidities were identified in 70% of the patients, with hypertension (70%), dyslipidemia (62%) and cardiovascular disease (30%) being the most common. Initial symptoms included fever (100%), coughing (85%), dyspnea (76%), diarrhea (42%) and asthenia (21%). At triage, the average of SpO_2_ was 89%.

**Table 1.** Baseline Characteristics of 48 Patients With COVID-19 at triage, prior hospitalization.

| Clinical Characteristics | All (N= 48) | Non-ICU (N= 27) | ICU (N= 21) | P value |
| --- | --- | --- | --- | --- |
| **At triage** |  |  |  |  |
| **Age, yrs.** | 65.98 (13.91)<br>[33-88] | 66.30 (14.90)<br>[33-88] | 65.57 (12.87)<br>[44-82] | 0.856 |
| **Males, n %** | 32 (67%) | 18 (67%) | 14 (67%) | 1 |
| **Fever, °C** | 37.03 (0.94)<br>[36-39] | 36.84 (0.88)<br>[36-39] | 37.28 (0.98)<br>[36-39] | 0.147 |
| **Systolic Pressure, mmHg** | 129.6 (18.9)<br>[90-180] | 130.7 (16.6)<br>[90-180] | 128.1(21.9)<br>[90-180] | 0.642 |
| **Diastolic Pressure, mmHg** | 73.3 (11.7)<br>[50-111] | 75.9 (12.1)<br>[50-110] | 70.03 (10.44)<br>[52-92] | 0.058 |
| **Heart Rate, bpm** | 85.6 (14.59)<br>[58-120] | 86.9(17.4)<br>[58-120] | 83.8 (11.2)<br>[58-106] | 0.712 |
| **SpO<sub>2</sub>, %** | 89.31(10.64) | 93.44(6.63) | 84(12.51) | **<0.001\*** |

|  | [38-99] | [66-99] | [38-99] |  |
| --- | --- | --- | --- | --- |
| **Symptoms reported n%** |  |  |  |  |
| **Asthenia** | 10/46(21%) | 6/25 (22%) | 4/21 (19.1%) | 1 |
| **Dyspnea** | 35/46 (76%) | 16/26 (61%) | 19/20 (95) | **0.022\*** |
| **Vomiting** | 6/47 (13%) | 4/25 (16%) | 2/21 (9%) | 0.870 |
| **Diarrhea** | 16/38 (42%) | 12/25 (44%) | 4/13 (31%) | 0.070 |
| **Coughing** | 39/46 (85%) | 20/27 (74%) | 19/20 (95%) | 0.225 |
| **Fever** | 48 (100%) | 27 (100%) | 21 (100%) | 1 |
| **ARDS** | 20/46 (44%) | 0/27 (0%) | 21/21 (100%) | **<0.001\*** |
| **Pneumonia** | 44/47 (94%) | 24/27 (89%) | 20/20 (100%) | 0.078 |
| **Bilateral pneumonia** | 44/47 (94%) | 21/27 (77%) | 20/20 (100%) | NA |
| **Comorbidities n%** |  |  |  |  |
| **Hypertension** | 33/47 (70%) | 22/27 (82%) | 11/20 (55%) | 0.101 |
| **Dyslipidemia** | 29/47 (62%) | 16/27 (60%) | 13/20 (65%) | 1 |
| **Type 2 Diabetes** | 11/45 (24%) | 9/27 (33%) | 5/20 (25%) | 0.286 |
| **Cardiovascular disease** | 14/47 (30%) | 7/27 (26%) | 7/20 (35%) | 0.726 |
| **Ictus** | 3/46 (6%) | 2/27 (7%) | 1/20 (5%) | 0.662 |
| **Cancer or another malignancy** | 10/47 (21%) | 4/27 (15%) | 6/20 (30%) | 0.640 |
| **COPD** | 5/47 (11%) | 4/27 (15%) | 1/20 (5%) | 0.544 |
| **VIH** | 1/46 (2%) | 0/26 (0) | 1/20 (5%) | 0.894 |
| **Renal chronic disease** | 8/46 (17%) | 4/27 (15%) | 4/19 (21%) | 0.877 |
| **Other, n%** | 26/47 (55%) | 13/27 (52%) | 12/20 (60%) | 0.921 |
| **Smoking** | 10/47 (21%) | 6/26 (22%) | 4/20 (19%) | 0.934 |

Data are mean (SD), range [ ] or %
\*statistically significant

ICU patients compared to non-ICU patients, presented a significantly lower SpO_2_ (84%±12.51 vs. 93%±6.63, *p* < .001), and a higher prevalence of dyspnea (95% vs 61%, *p* = .022) and ARDS (100% vs 0%, p < .001).

An abnormal chest radiograph presenting bilateral pneumonia was observed in 44 patients (94%) at admission. Only 3 non-ICU patients (11%) had unilateral pneumonia. Representative lung images showing interstitial lung abnormalities of the 8 deceased ICU patients are presented in Figure 1. As of April 27, 2020, the overall mortality was 21% (10/48). 8 patients died in the ICU group (38%) vs. 2 in non-ICU (0.07%) (Figure 2).

**Figure 1.**
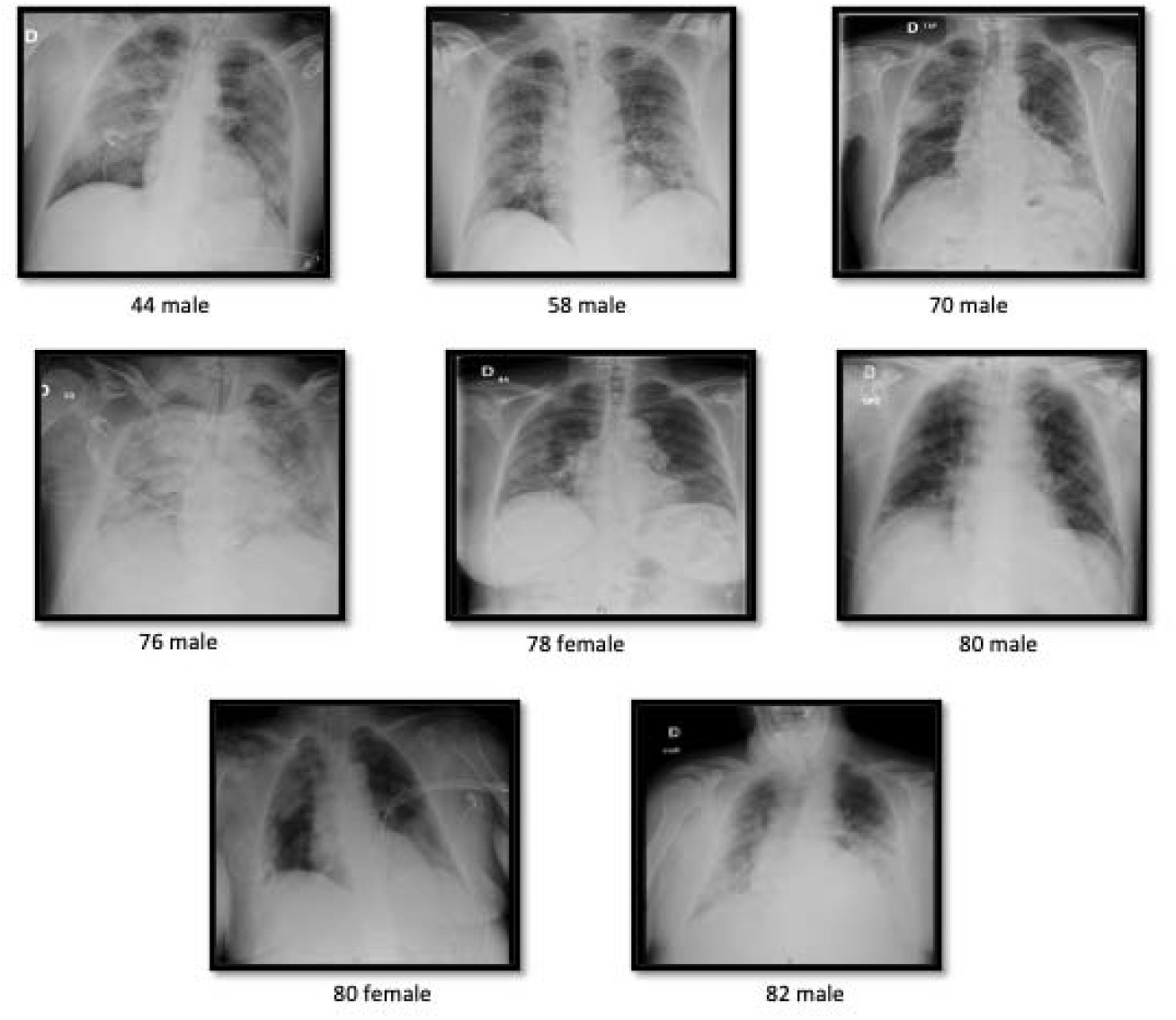
Chest x-ray images from all the deceased ICU patients. Most of the patients presented bilateral pneumonia at triage.

**Figure 2.**
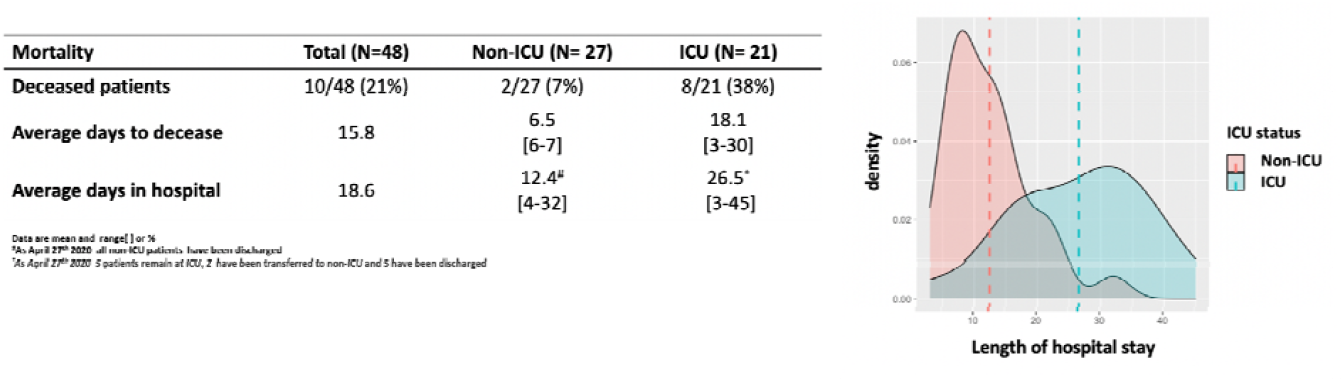
Mortality and length of hospital stay of the 48 COVID-19 patients. A) Mortality, average days to decease and days in the hospital in non-ICU and ICU patients with COVID-19 in Mallorca, B) Density plot of the length of hospital stay showing the average days in the hospital red non ICU patients and green ICU patients with COVID-19

Treatments used to treat COVID-19 patients are summarized in Supplemental Table 1. During hospitalization, 100 % of the patients received Hidroxychloroquine or Chloroquine, 98% received Kaletra, an antiviral treatment combining lopinavir and ritonavir, 100% received antibiotics, 54% corticosteroids and 100% received oxygen therapy. Tocilicumab was used in 48% of the ICU patients. We only had information on corticosteroids treatment in ICU. Due to missing information on renal treatment those data are not presented. Invasive mechanical ventilation was received in all ICU patients.

### Laboratory findings

#### Hematologic measures

Table 2 shows the hematologic measures at admission (mean (SD), range [] or %). Compared with the normal range, leukocyte or white blood cell (WBC) counts were normal in non-ICU vs. ICU patients, whereas lymphocyte counts were significantly lower in ICU patients (1.03 vs. 0.7 ×10^9^/L respectively, *p*= .002). and below normal range ([1.00 − 4.5 × 10^9^/L]). Similarly, monocytes counts were significantly lower in the ICU group compared to non-ICU (0.58 vs. 0.40 × 10^9^/L, p= .029). After adjusting for possible confounders such as age, sex, race, smoking, spO2, hypertension and dyslipidemia, lymphocyte and monocytes counts remained significantly lower in ICU patients (*p* =.045 and p= 0.040 respectively). When adding comorbidities such as type 2 diabetes, cardiovascular diseases, and COPD into the model, the differences in monocyte and lymphocyte counts disappeared, although they were close to significant (*p*=.055 and *p*= .158 respectively). No differences in the number of red blood cells, hematocrit, hemoglobin, mean corpuscular volume (MCV), mean corpuscular hemoglobin (MCH), red blood cell distribution width (RDW), platelet distribution width (PDW), platelets and mean platelet volume (MVP) were found between non-ICU and ICU patients.

**Table 2.**
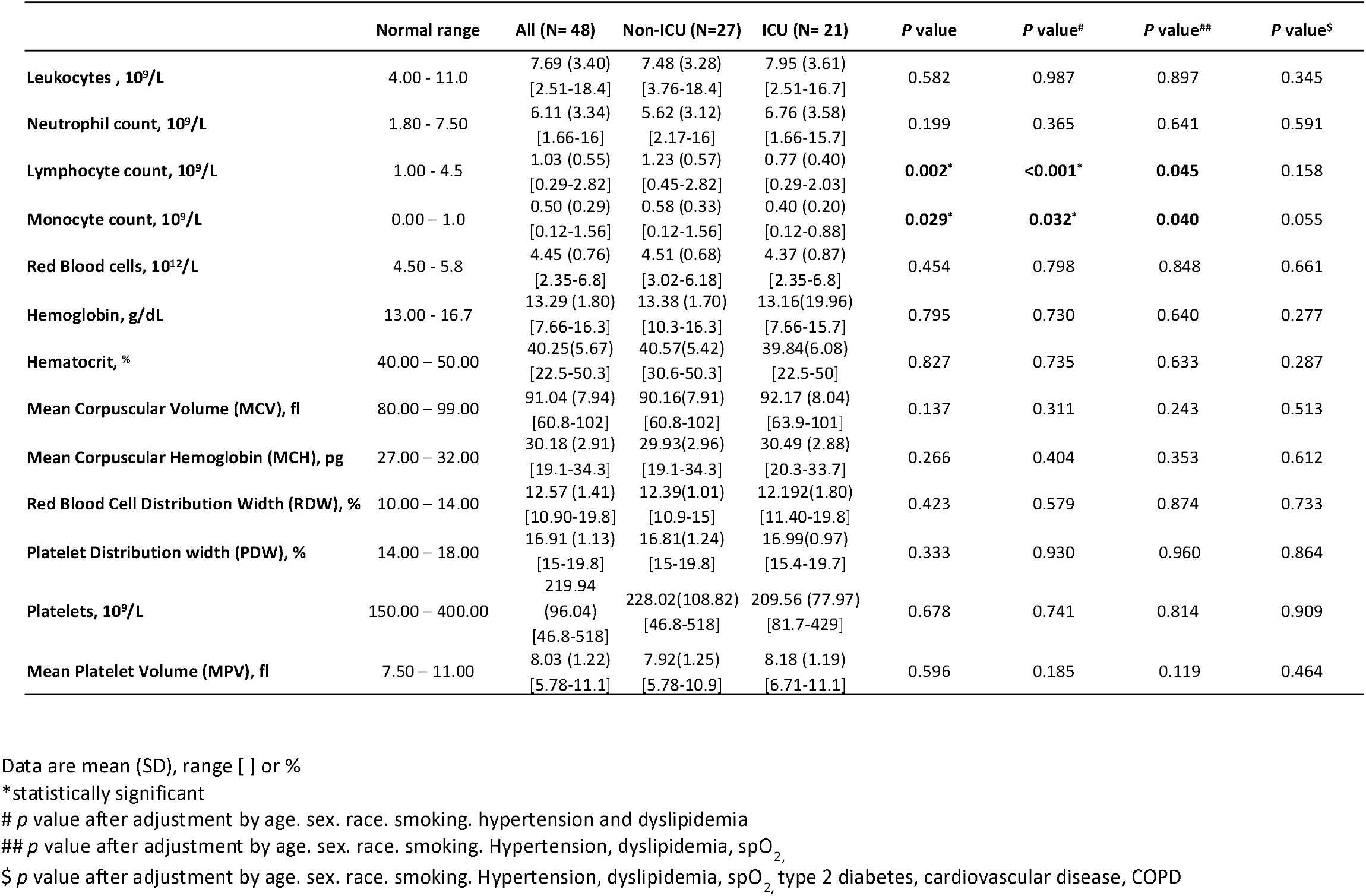
Baseline Laboratory Measures of 48 Patients With COVID-19 at triage, prior hospitalization.

| | Normal range | All (N= 48) | Non-ICU (N=27) | ICU (N= 21) | P value | P value<sup>#</sup> | P value<sup>##</sup> | P value<sup>\$</sup> |
| --- | --- | --- | --- | --- | --- | --- | --- | --- |
| Leukocytes , 10<sup>9</sup>/L | 4.00 - 11.0 | 7.69 (3.40)<br>[2.51-18.4] | 7.48 (3.28)<br>[3.76-18.4] | 7.95 (3.61)<br>[2.51-16.7] | 0.582 | 0.987 | 0.897 | 0.345 |
| Neutrophil count, 10<sup>9</sup>/L | 1.80 - 7.50 | 6.11 (3.34)<br>[1.66-16] | 5.62 (3.12)<br>[2.17-16] | 6.76 (3.58)<br>[1.66-15.7] | 0.199 | 0.365 | 0.641 | 0.591 |
| Lymphocyte count, 10<sup>9</sup>/L | 1.00 - 4.5 | 1.03 (0.55)<br>[0.29-2.82] | 1.23 (0.57)<br>[0.45-2.82] | 0.77 (0.40)<br>[0.29-2.03] | **0.002\*** | **<0.001\*** | **0.045** | 0.158 |
| Monocyte count, 10<sup>9</sup>/L | 0.00 – 1.0 | 0.50 (0.29)<br>[0.12-1.56] | 0.58 (0.33)<br>[0.12-1.56] | 0.40 (0.20)<br>[0.12-0.88] | **0.029\*** | **0.032\*** | **0.040** | 0.055 |
| Red Blood cells, 10<sup>12</sup>/L | 4.50 - 5.8 | 4.45 (0.76)<br>[2.35-6.8] | 4.51 (0.68)<br>[3.02-6.18] | 4.37 (0.87)<br>[2.35-6.8] | 0.454 | 0.798 | 0.848 | 0.661 |
| Hemoglobin, g/dL | 13.00 - 16.7 | 13.29 (1.80)<br>[7.66-16.3] | 13.38 (1.70)<br>[10.3-16.3] | 13.16(19.96)<br>[7.66-15.7] | 0.795 | 0.730 | 0.640 | 0.277 |
| Hematocrit, % | 40.00 – 50.00 | 40.25(5.67)<br>[22.5-50.3] | 40.57(5.42)<br>[30.6-50.3] | 39.84(6.08)<br>[22.5-50] | 0.827 | 0.735 | 0.633 | 0.287 |
| Mean Corpuscular Volume (MCV), fl | 80.00 – 99.00 | 91.04 (7.94)<br>[60.8-102] | 90.16(7.91)<br>[60.8-102] | 92.17 (8.04)<br>[63.9-101] | 0.137 | 0.311 | 0.243 | 0.513 |
| Mean Corpuscular Hemoglobin (MCH), pg | 27.00 – 32.00 | 30.18 (2.91)<br>[19.1-34.3] | 29.93(2.96)<br>[19.1-34.3] | 30.49 (2.88)<br>[20.3-33.7] | 0.266 | 0.404 | 0.353 | 0.612 |
| Red Blood Cell Distribution Width (RDW), % | 10.00 – 14.00 | 12.57 (1.41)<br>[10.90-19.8] | 12.39(1.01)<br>[10.9-15] | 12.192(1.80)<br>[11.40-19.8] | 0.423 | 0.579 | 0.874 | 0.733 |
| Platelet Distribution width (PDW), % | 14.00 – 18.00 | 16.91 (1.13)<br>[15-19.8] | 16.81(1.24)<br>[15-19.8] | 16.99(0.97)<br>[15.4-19.7] | 0.333 | 0.930 | 0.960 | 0.864 |
| Platelets, 10<sup>9</sup>/L | 150.00 – 400.00 | 219.94<br>(96.04)<br>[46.8-518] | 228.02(108.82)<br>[46.8-518] | 209.56 (77.97)<br>[81.7-429] | 0.678 | 0.741 | 0.814 | 0.909 |
| Mean Platelet Volume (MPV), fl | 7.50 – 11.00 | 8.03 (1.22)<br>[5.78-11.1] | 7.92(1.25)<br>[5.78-10.9] | 8.18 (1.19)<br>[6.71-11.1] | 0.596 | 0.185 | 0.119 | 0.464 |

Data are mean (SD), range [ ] or %
\*statistically significant

#### Coagulation function, biochemical and inflammation measures

Table 3 shows the coagulation function, biochemical and inflammation measures admission (mean (SD), range [] or %). Due to the outliers in D-dimer and Ferritin, we show the data in these measures in mean (SD), range [], median (IQR), and in log scale. Prothrombin time was slightly higher in ICU compared to non-ICU patients (p = .038), but when adjusted for age, sex, race, smoking, SpO2, hypertension, dyslipidemia type 2 diabetes, cardiovascular diseases, and COPD, the difference disappeared. Mean levels of fibrinogen (713.63 vs. 200-500 md/dL) and median D-dimer (358 n vs. 0.00-255 ng/mL) were over normal range in COVID-19 patients. No differences were observed in fibrinogen or D-dimer levels among groups, but after adjusting for possible confounders, D-dimer levels were significant (*p*= .020).

**Table 3.** Baseline coagulation function, biochemical and inflammation measures.

| | Normal range | All (N= 48) | Non-ICU (N= 27) | ICU (N= 21) | P value | P value<sup>#</sup> | P value<sup>\$</sup> |
| --- | --- | --- | --- | --- | --- | --- | --- |
| Prothrombin time, s | 8.5 - 15.0 | 13.70 (1.59)<br>[11.4-20.2] | 13.20(1.12)<br>[11.40-15.50] | 14.33(1.89)<br>[11.9-20.2] | **0.038\*** | 0.614 | 0.461 |
| Fibrinogen, mg/dL | 200 - 500 | 713.63 (160.50)<br>[405-1185] | 686.44(151.77)<br>[420-1118] | 748.57 (168.27)<br>[405-1185] | 0.212 | 0.093 | 0.078 |
| D-Dimer, ng/mL* | 0.00 - 255 | 1745.08 (6495.60) / 358 (262-609)*<br>[94-44808] | 2405.93(8583.03)/ 358 (201-2406)*<br>[150-44808] | 895.43(1427.10)/ 350 (272-895)*<br>[94-5105] | 0.731 | 0.279 | **0.020\*** |
| Log (D-Dimer) |  | 6.17(1.17)<br>[4.5-10.7] | 6.18(1.32)<br>[5.0-10.7] | 6.16(1.0)<br>[4.5-8.5] | 0.967 | 0.412 | 0.121 |
| Creatinine, mg/dL | 0.72 - 1.25 | 1.10 (0.83)<br>[0.6-5.4] | 1.09(0.68)<br>[0.64-3.82] | 1.11(1.01)<br>[0.57-5.40] | 0.739 | 0.842 | 0.405 |
| Glomerular Filtration Rate, mL/min |  | 77.17(25.44)<br>[11-125] | 76.15(25.99)<br>[16-116] | 78.48(25.28)<br>[11-125] | 0.830 | 0.852 | 0.883 |
| Ferritin, ng/mL | 20 - 274 | 2074.37 (5903)/ 572 (279-1401)*<br>[76.9-40000] | 750.58(1374.38)/352(172-652)*<br>[76.9-7245.9] | 3776.39(8603.70)/ 1373(571-2598)*<br>[261.8-40000] | **<0.001\*** | 0.054 | 0.084 |
| Log(Ferritin) |  | 6.51(1.32)<br>[4.3-10.6] | 5.93(1.09)<br>[4.3-8.8] | 7.26(1.24)<br>[5.57-10.6] | **<0.001** | **<0.001\*** | **0.008\*** |
| Total Bilirubin, mg/dL | 0.2 - 1.2 | 0.79 (0.45)<br>[0.3-22] | 0.70(0.28)<br>[0.34-1.43] | 0.91(0.58)<br>[0.41-2.20] | 0.339 | 0.399 | 0.387 |
| Albumin, g/dL | 3.50 - 5.50 | 3.47 (0.69)<br>[1.8-4.7] | 3.92(0.42)<br>[3.36-4.74] | 2.90(0.52)<br>[1.84-3.64] | **<0.001\*** | **<0.001\*** | **<0.001** |
| Aspartate aminotransferase (AST), U/L | 5.0-34.0 | 44.50(27.66)<br>[3-125] | 40.41(30.15)<br>[3-118] | 49.76(23.77)<br>[22-125] | **0.040\*** | 0.062 | 0.369 |
| Alanine aminotransferase (ALT), U/L | 1.0-55.0 | 39.75(30.28)<br>[9-121] | 40.67(35.23)<br>[9-121] | 38.57(23.19)<br>[14-94] | 0.323 | 0.367 | 0.665 |
| Phosphatase, U/L | 40 - 150 | 80.88(45.19)<br>[25-250] | 74.96(27.59)<br>[34-143] | 88.48(60.83)<br>[25-250] | 0.909 | 0.890 | 0.311 |
| Gamma-glutamyl transferase (GGT), U/L | 12.0-64.0 | 79.60(80.77)<br>[11-433] | 69.04(51.16)<br>[13-186] | 93.19(106.99)<br>[11-433] | 0.843 | 0.338 | 0.227 |
| Lactate dehydrogenase (LDH), U/L | 125 - 243 | 479.04(548.36)<br>[156-4038] | 335.82(123.6)<br>[156-545] | 663.19 (789.6)<br>[237-4038] | **<0.001\*** | **0.005\*** | **0.113** |
| Creatine Kinase (CK), U/L | 30 - 200 | 181.79(298.45)<br>[27-1909] | 202.89(375.21)<br>[29-1909] | 154.67(157.89)<br>[27-736] | 0.473 | 0.321 | 0.265 |
| Log(CK) |  | 4.64(0.95)<br>[3.3-7.5] | 4.60(1.04)<br>[3.37-7.55] | 4.69(0.84)<br>[3.3-6.0] | 0.744 | 0.601 | 0.536 |
| Triglycerides, mg/dL | 0 - 150 | 173.79 (74.46)<br>[64-399] | 163.22(68.40)<br>[64-388] | 187.38(81.26)<br>[79-399] | 0.284 | 0.215 | 0.651 |
| Glucose, mg/dL | 70 - 110 | 131.23 (59.18)<br>[80-431] | 131.78(65.38)<br>[80-431] | 130.52(51.69)<br>[89-337] | 0.868 | 0.601 | 0.347 |
| Urea, mg/dL | 15 - 50 | 47.46 (38.64)<br>[14-193] | 51.04(40.51)<br>[16-176] | 42.86(36.55)<br>[14-193] | 0.442 | 0.423 | **0.045\*** |
| Troponin I, ng/L | 0.00 - 34.0 | 210.76(1094.37)<br>[1.9-7572.6] | 76.01(184.38)<br>[1.90-723.1] | 384.00(1647.5)<br>[1.9-7572.6] | 0.412 | 0.475 | **0.034\*** |
| C reactive protein (CRP), mg/L | 0.0 - 5.0 | 150.43(96.62)<br>[0.5-350.4] | 101.30(73.27)<br>[0.5-238.9] | 213.59(86.68)<br>[98.80-350.4] | **<0.001\*** | **0.011\*** | **0.028** |
| Procalcitonin, ng/mL | 0.00 - 0.05 | 1.02(4.98)<br>[0.01-34.7] | 0.10(0.11)<br>[0.01-0.43] | 2.21(7.47)<br>[0.02-34.66] | **<0.001\*** | **<0.001\*** | **0.016** |

Data are mean (SD), range [ ] or %
\*statistically significant

All the biochemical measures were in normal range among groups except ferritin (572 ng/dL vs. 20-274 ng/dL), aspartate aminotransferase (AST) (44.5 U/L vs. 5.0-34.0 U/L), glucose (131.2mg/dL vs. 70-100 mg/dL), and triglycerides (173.8 mg/dL vs. 0-150 mg/dL), where the overall mean was above the normal range.

No differences in creatinine, glomerular filtration rate, total bilirubin, alanine aminotransferase (ALT), phosphatase, gamma-glutamyl transferase (GGT), creatine kinase (CK), triglycerides, glucose, or urea were found among groups. Urea was significantly lower in ICU patients when adjusted by age, sex, race, smoking, SpO_2_, hypertension, dyslipidemia type 2 diabetes, cardiovascular diseases, and COPD (*p =* .045). AST was significantly higher in ICU patients (*p*= .040), although after adjusting for all the confounders, the difference disappeared. Interestingly, albumin, was significantly lower in ICU patients (p<.001), even in adjusted models (p<.001). After this finding, we decided to study the association of albumin with outcomes of prognosis of the disease. Overall, we found that lower albumin levels were associated with longer hospital length and mortality (r= −0.472, p <.001 and r= −0.424, p=.003, respectively) in COVID-19 patients (Figure 3). Albumin levels were also positive correlated with absolute number of lymphocytes (r= 0.368, p <0.001), and negative correlated with inflammatory markers such as procalcitonin (r= −0.555, p <.001), LDH (r= −0.443, p=.002), CRP (r= −0.390, p=0.006), troponin I (r= −0.321, p=.026), ferritin (r= −0.506, p<0.001) and MCV (r= −0.424, p= 0.003) among COVID-19 patients.

**Figure 3.**
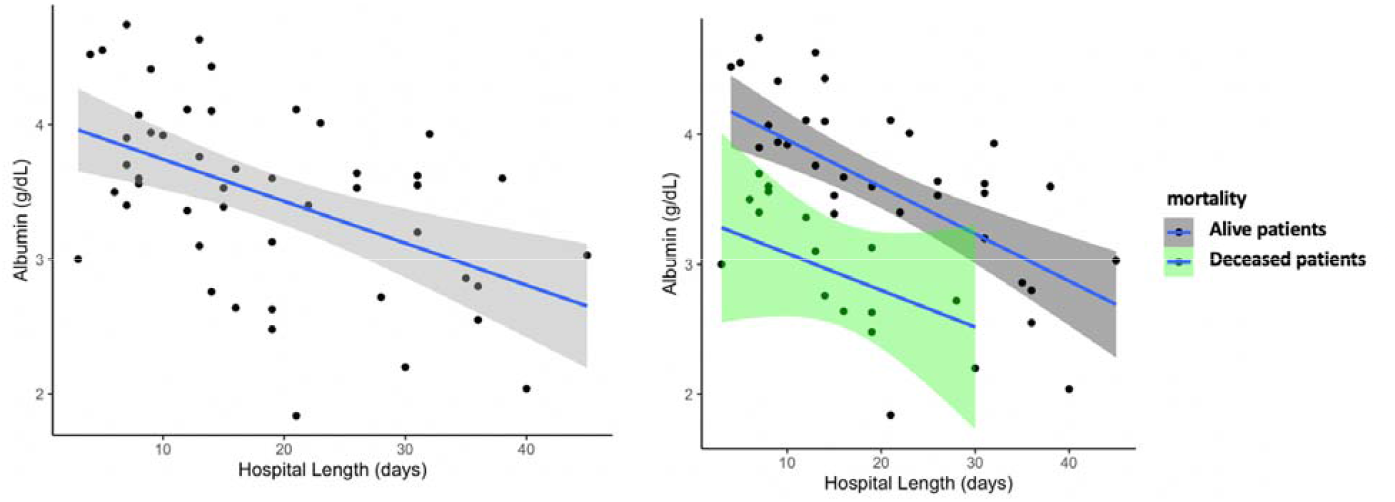
Association of serum albumin levels with hospital length in COVID-19. Patients with lower albumin stayed longer days in the hospital. On the right panel, scatterplot showing the association of albumin levels and hospital day by groups, in green the deceased patients (10 patients) and in grey the alive patients (38 patients)

Interestingly, COVID-19 patients who died presented higher MCV compared to COVID-19 who survived (mean vs 95.80±3.32 vs 89.79±8.35 fl respectively, p= 0.001) (data not shown).

Finally, the mean of each inflammatory marker was in range among COVID-19 patients except lactate dehydrogenase (LDH) (479 vs. 124-243 U/L), troponin I (210.76 vs. 0.0-34.0 ng/L), C reactive protein (CRP) (150.43 vs. 0.0-5.0 mg/L), and procalcitonin (1.02 vs. 0.0-0.05 ng/mL), which were all higher. LDH, CRP, and procalcitonin levels were significantly higher in ICU patients compared to non-ICU (p < .001). Troponin I levels became statistically different after adjusting for age, sex, race, smoking, spO2, hypertension, dyslipidemia type 2 diabetes, cardiovascular diseases, and COPD (p= .045). Figure 4 shows the boxplots of the laboratory measures that presented more differences among non-ICU and ICU patients with COVID-19.

**Figure 4.**
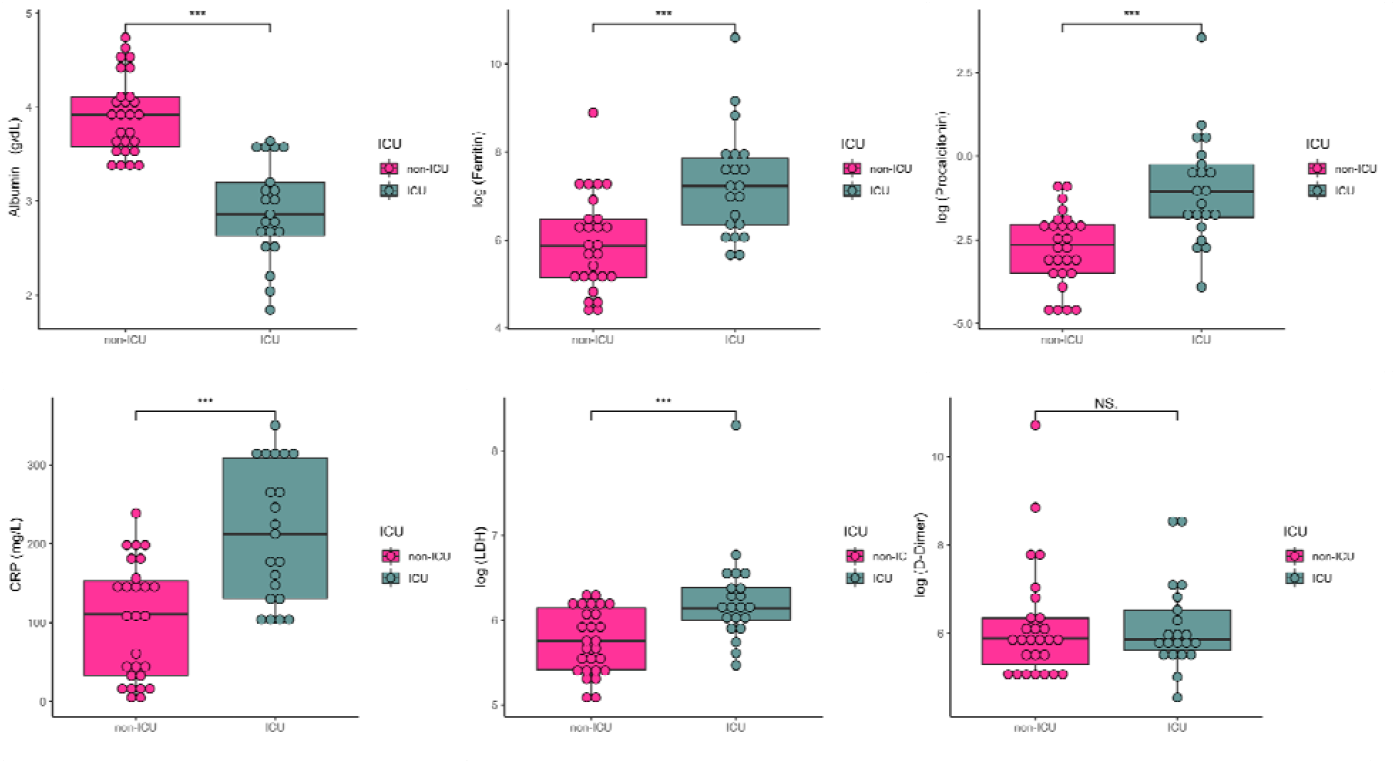
Representative Scatterplots of inflammatory markers in the 48 patients with COVID-19.

## Discussion

To our knowledge, this study represents the first case series of sequentially hospitalized adult patients with confirmed COVID-19 in Spain. Knowledge of the baseline characteristics and outcomes of critically ill patients is crucial for health and government officials to address local outbreaks. Efforts are being made in order to fully characterize the clinical characteristics of COVID-19 nationwide. The overall mortality in this case series was 21%. Studies with higher numbers of patients are likely to find lower mortality rates.

The majority of patients in this case series were hospitalized because of low SpO_2_ (89%), and presentation of bilateral pneumonia (94%). 100% of the ICU patients, presented ARDS. These data are similar to those shown by similar studies from Italy and China. Most of the patients reported fever and a cough one week prior to hospitalization. Interestingly, among the 48 patients, 44% reported diarrhea as well, and this percentage was higher in the non-ICU group (44% vs 31%, *p* = .070). These numbers are higher to those shown in previous reports (17-20%). Other common symptoms at onset of illness were fever, dry cough, and dyspnea. 100% of the patients required oxygen therapy. All ICU patients required mechanical ventilation.

Early publications from other countries show that epidemiological features such as age, sex and comorbidities play an important role in disease progression.^15,16^ Men are more likely to show a poor prognosis than women.^15^ Diabetes, chronic obstructive pulmonary disease (COPD), and hypertension are often comorbid in severe COVID-19.^16-18^ In our COVID-19 patients, hypertension was the most prevalent disease (70%), followed by dyslipidemia, cardiovascular disease and diabetes.

Overall, ICU patients presented lymphopenia and normal WBC, similar to other smaller case series reports of critically ill patients in the US, China, Singapore or Italy.^1,19-22,^ Patients who died, also presented lower lymphocytes compared to survivors (p<.0001, data not shown). Interestingly, MCV was also lower in deceased patients, although the numbers were in range (data not shown). We also found that critically patients had hypoalbuminemia, and that this remained statistically significant even after adjusting for possible confounders. To our knowledge, only two studies have reported similar data on albumin levels in COVID19, but these results have not been emphasized^5,23^. Previous research has shown that hypoalbuminemia is a strong predictor of 30-day, all-cause mortality in critically ill patients.^24^ In fact, in our case series lower albumin levels were associated with mortality and length of stay (Figure 3). Serum albumin levels on admission predicts the need of intensive respiratory support in adult patients with influenza A (H1N1).^25^ Lower levels of albumin have also been associated with higher inflammation, hypercoagulation, and carotid atherosclerosis in people with human immunodeficiency virus infection (HIV).^26^ Interestingly, a recent report found that treatment with chemically modified albumin could confers protection against the entry of the Ebola virus in cells.^27^ Albumin levels might be involved in poor outcomes in COVID-19 patients as well. Similar to other studies, ICU patients presented with inflammatory markers above the normal range, especially D-dimer, LDH, ferritin, fibrinogen, CPR, troponin I and procalcitonin. Contrary to other studies, the levels of D-dimer did not differ among groups. D-dimer levels were statistically significant only when the data were adjusted for sex, age, race, smoking, SpO_2_ and comorbidities. Higher levels of these inflammatory markers are indicative of coagulation, cardiac and renal dysfunction.

Pro-inflammatory factors play a central role in COVID-19 severity, especially in patients with comorbidities.^6,10^ The rapid virus replication rate along with interferon attenuation mechanisms and the accumulation of macrophages in the lungs seem to be important triggers of this “cytokine storm”.^11^ We believe that it is imperative to measure inflammatory cytokines involved in the cytokine storm that has been seen in COVID-19. Polymorphisms in the angiotensin-converting enzyme receptor 2 (ACE2) are also likely to be involved in poor outcomes.^28-30^

There is an urgent need to improve our understanding on the phenotype profiles behind the progress from mild to severe or critical COVID-19. To date, there is no specific treatment against COVID-19. In our cases series, 100% of the patients received a combination of antibiotic and antiviral treatments, and 81% received an antimalarial drug after being admitted to the hospital. Administering immunomodulators aimed to reduce COVID-19-driven inflammation comes with serious risks. Further studies are needed to characterize the effects of these drugs in cardiovascular, renal and pulmonary function in COVID-19 patients.

### Limitations of the study

This study has several limitations. First, the study population is small and only includes patients from Son Llatzer hospital, one of the main hospitals within Balearic Health system. There may be a selection bias when identifying factors that differ between non-ICU and ICU patients, even though the results were adjusted for known confounders, including age, sex, race, SpO_2_, smoking status and comorbidities. Also, this study is the first to show the characteristics of severe and critically ill COVID-19 adult patients in Spain. Secondly, this is a retrospective study done in an emergency situation. Third, the follow-up data could not be analyzed due to a delay in the update of the electronic health record database. Finally, some patients presented elevated biomarker data in some laboratory measures, and we did not exclude them from the analysis due to the small sample size and because those numbers were physiological and not due to technical error.

## Conclusion

In conclusion, critically ill patients with COVID-19 present at triage lymphopenia, hypoalbuminemia as well high levels of inflammation. Lower levels of albumin were associated with poorer outcomes in COVID-19 patients. Albumin might be of importance because of its association with disease severity in patients infected with SARS-CoV-2. This small case series provides the first steps towards a comprehensive clinical characterization of severe and critical COVID-19 adult patients in Spain.

## Data Availability

The data including in this manuscript will be available prior request to the main and/or corresponding author

## Dissemination to participants and related patient and public communities

No study participants were involved in the preparation of this article. The results of the article will be summarized in media press releases from IdISBa and presented at relevant conferences.

## Disclaimer

The views expressed in this article are those of the authors and do not represent the views of the Health Research Institute of the Balearic Islands, Son Llatzer University Hospital, or any other government entity.

## Author contributions

A.M., A.C., A.S. acquired the data. M.G. analyzed data. R.R. and M.G. supervised the project and wrote the manuscript. All authors participated in scientific discussions.

## TABLE LEGENDS

**Supplemental Table 1.**
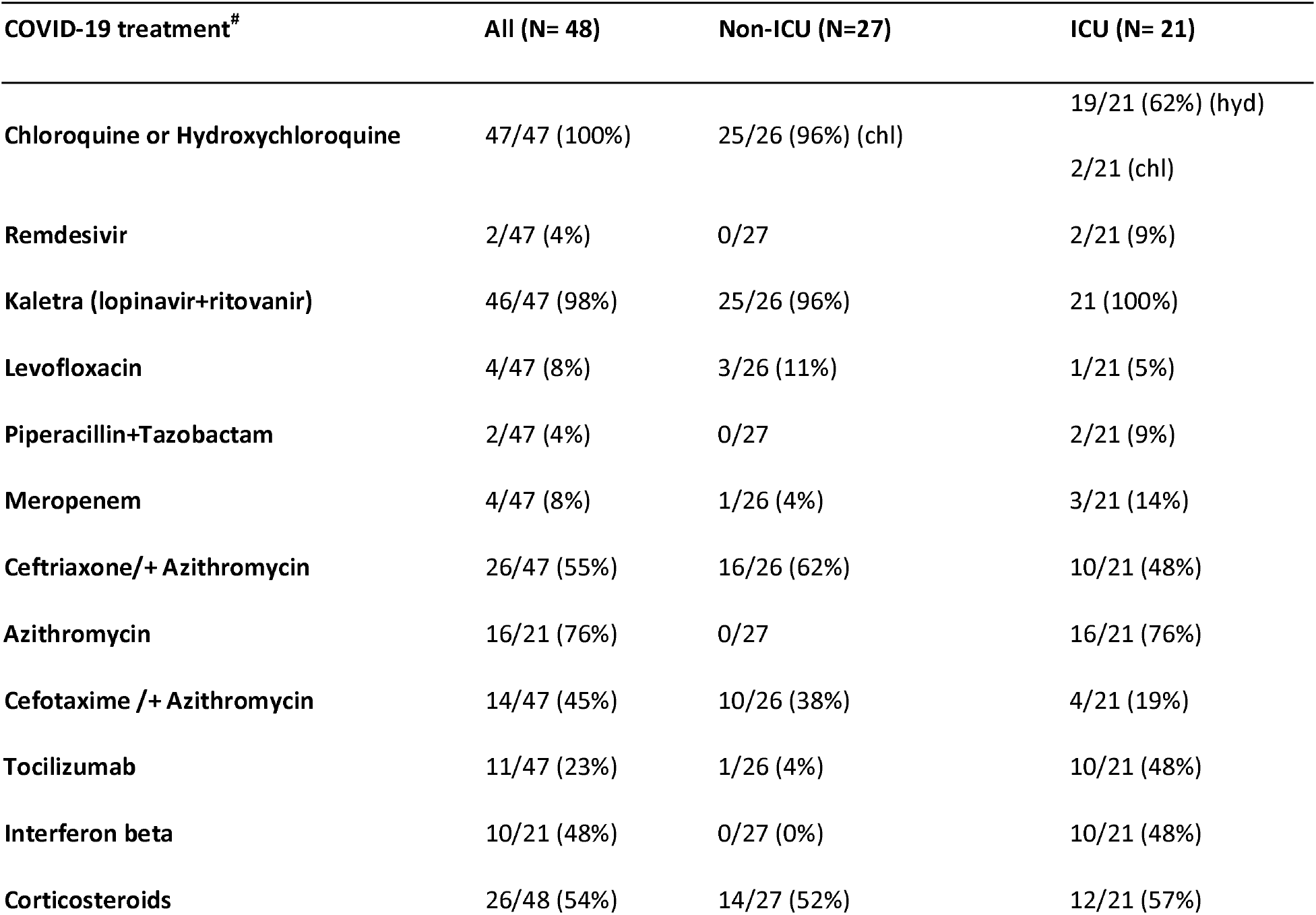
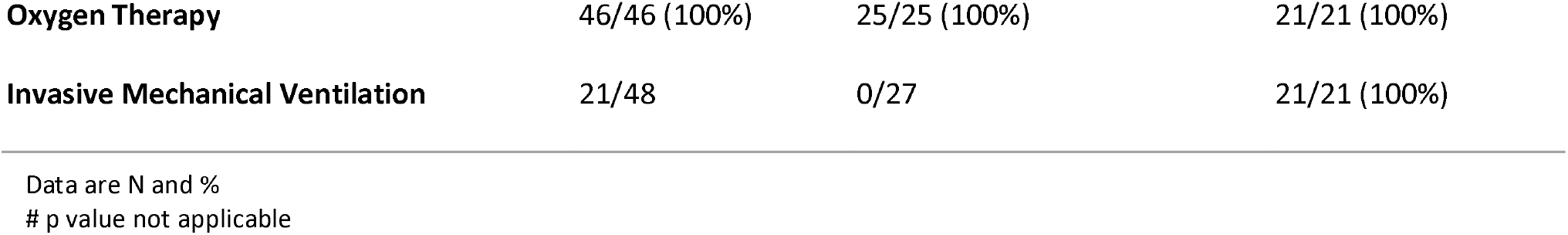
Treatments used in the 48 COVID-19 patients during hospitalization.

| COVID-19 treatment<sup>#</sup> | All (N= 48) | Non-ICU (N=27) | ICU (N= 21) |
| --- | --- | --- | --- |
| Chloroquine or Hydroxychloroquine | 47/47 (100%) | 25/26 (96%) (chl) | 19/21 (62%) (hyd)<br>2/21 (chl) |
| Remdesivir | 2/47 (4%) | 0/27 | 2/21 (9%) |
| Kaletra (lopinavir+ritovanir) | 46/47 (98%) | 25/26 (96%) | 21 (100%) |
| Levofloxacin | 4/47 (8%) | 3/26 (11%) | 1/21 (5%) |
| Piperacillin+Tazobactam | 2/47 (4%) | 0/27 | 2/21 (9%) |
| Meropenem | 4/47 (8%) | 1/26 (4%) | 3/21 (14%) |
| Ceftriaxone/+ Azithromycin | 26/47 (55%) | 16/26 (62%) | 10/21 (48%) |
| Azithromycin | 16/21 (76%) | 0/27 | 16/21 (76%) |
| Cefotaxime /+ Azithromycin | 14/47 (45%) | 10/26 (38%) | 4/21 (19%) |
| Tocilizumab | 11/47 (23%) | 1/26 (4%) | 10/21 (48%) |
| Interferon beta | 10/21 (48%) | 0/27 (0%) | 10/21 (48%) |
| Corticosteroids | 26/48 (54%) | 14/27 (52%) | 12/21 (57%) |
| **Oxygen Therapy** | 46/46 (100%) | 25/25 (100%) | 21/21 (100%) |
| **Invasive Mechanical Ventilation** | 21/48 | 0/27 | 21/21 (100%) |

Data are N and %

